# Longitudinal symptom and clinical outcome analysis of hospitalized COVID-19 patients

**DOI:** 10.1101/2022.01.11.22268908

**Authors:** Arturas Ziemys

## Abstract

COVID-19 pandemics increased patient hospitalization impacting the hospital operations and patient care beyond COVID-19 patients. Although longitudinal symptom analysis may provide prognostic utility about clinical outcomes and critical hospitalization events of COVID-19 patients, such analysis is still missing. Here, we have analyzed over 10,000 hospitalized COVID-19 patients in the Houston Methodist Hospital at the Texas Medical Center from the beginning of pandemics till April of 2020. Our study used statistical and regression analysis over symptoms grouped into symptom groups based on their anatomical locations. Symptom intensity analysis indicated that symptoms peaked at the time of admission and subsided within the first week of hospitalization for most of the patients. Patients surviving the infection (n=9,263), had faster remission rates, usually within the first days of hospitalization compared to sustained symptom for the deceased patient group (n=1,042). The latter had also a longer hospitalization stay and more comorbidities including diabetes, cardiovascular, and kidney disease. Inflammation-associated systemic symptoms (Systemic) such as fever and chills, and lower respiratory system specific symptoms (Lower Respiratory System) such as shortness of breath and pneumonia, were the most informative for the analysis of longitudinal symptom dynamics. Our results suggest that the symptom remission rate could possess prognostic utility in evaluating patient hospitalization stay and clinical outcomes early in hospitalization. We believe knowledge and information about symptom remission rates can be used to improve hospital operations and patient care by using common and relatively easy to process source of information.

## Introduction

The outbreak of COVID-19 pandemics has resulted in a large number of patients infected, hospitalized, and deceased. The infection stressed hospital operations because a large influx of COVID19 patients saturated capacities of intensive care and exhausted other resources disrupting hospital operations affecting not only COVID-19 patients, but also patients with other acute or chronic conditions [1-3]. The pandemic has prompted attention to the optimization of care [4]. Abilities to acquire prognostic information on the outcomes of hospitalized COVID-19 patients could be of high benefit when seeking to improve clinical outcomes by optimizing decisions and operations in patient care.

To date, the success of experimental therapeutic treatments was marginal and focused on symptom alleviation therapy rather than curing the viral infection [5]. Thus, long patient hospitalizations for patient life support are inevitable when treatment options are limited or absent during pandemics like COVID-19. However, hospitals collect large informative data about patients during their stay which can provide feedback to tailor patient care. Symptom recording is one of the fundamental care operations taking place in most healthcare environments and offering rich information for prognostic analysis. The symptomatology of the COVID-19 infection has been described well over the last year [6-8], with fever, cough, shortness of breath, or headache being among the most frequently observed symptoms. Efforts have been made to develop Artificial Intelligence (AI) approaches to analyze patient symptoms to empower clinical decision, as well [9, 10].

Longitudinal symptom analysis was less frequently approached to date, despite its potential to offer a deeper insight into patient wellbeing and the response to infections. Instead of having only the factual static approach for a specific symptom, the longitudinal analysis provides symptom remission dynamics over time. We believe the change in the symptom burden may provide quantitative metrics on how well patient’s immune and other systems respond to an infection, delivering insights well beyond the fact of the symptom presence. Few studies have approached such longitudinal analyses of COVID-19 patients so far possibly because those types of analyses are more involving and need more data. A large study (n=206,377) was conducted by analyzing primary-care electronic records and nationwide distributed surveys to analyze symptom dynamics in patient records with confirmed COVID-19 positive (n=2471) and negative individuals in Israel [11]. The study investigated the peak of symptoms in relation to the time of a positive test was confirmed, and the follow-up remission. The loss of taste and smell, weeks prior to a positive test, was found as the most discriminating factor for COVID-19 positive patients, whereas the onset of other specific symptoms occurred at different times in a patient. Furthermore, the study has found that self-reported symptoms method was more sensitive in capturing symptom dynamics. Other study conducted in the Unites States analyzed symptoms of over 500,000 users through self-reported surveys from April to May of 2020 [12]. This study has also found symptoms peaking at the time of positive COVID-19 test with most of the symptom remissions occurring within the first week after testing positive. This large population-based study has found that the analysis of self-reported symptoms was able to develop predictive models for COVID-19. Authors concluded that results of the symptom analysis could be applied to prioritize patient groups or triage patients. Longitudinal COVID-19 symptom onset was also approached with different mathematical modeling approaches with great potential for patient screening purposes [13].

Currently, only a limited number of studies performed longitudinal studies, most of the studies in COVID-19 patients were diffused offering snapshots of the symptom development, at most. Therefore, in-depth longitudinal symptoms analyses of hospitalized COVID-19 patients are critical to produce new decision therapeutic criteria. Here, we analyzed the symptom remission dynamics of COVID-19 patients hospitalized in the Houston Methodist Hospital at the Texas Medical Center. Our primary focus was to understand how clinical outcomes can be related to different symptom characters and to determine the specific symptom information that could be useful to predict patient outcomes or the length of hospitalization. We believe that such a study can provide benefits to patient care and hospital operations optimization.

## Material and methods

### Data source

The study protocol was approved by the Houston Methodist Hospital (HMH) review panel. The deidentified data set was acquired from the CURATOR data base in the HMH (PRO00025445). Only adult patients tested positive for COVID-19 were included. The ages of patients older than 90 year were capped at 90 years for deidentification purposes. Time records associated with patient were offset by the admission time.

### Symptom anatomical aggregation

Approximately 40 unique symptoms were extracted from records originating from patient flowsheets. To make the analysis more robust, we have aggregates symptoms based on their anatomical associations. Table 1 presents symptom groups, their abbreviations, and unique symptoms attributed to symptom groups. The symptom grouping is unique so that each individual symptom belongs to one symptom group only. The stiff neck symptom was attributed to the Central Nervous System (CNS) group because other viral brain infections, like viral meningitis [14, 15], possess such a symptom. Eight unique symptom groups were created. Symptom remission was defined as a rate of symptom frequency change over time within a specific group.

**Table 1.** Anatomical grouping of symptoms.

| Symptom group | Abbreviation | Symptoms |
| --- | --- | --- |
| Systemic | SYS | Chills and sweats; Chills/Repeated shaking with chills; Fever; Fever > 1 month; Loss of appetite; Night sweats; Unexplained weight loss |
| Upper respiratory system | URS | Nasal congestion |
| Lower Respiratory System | LRS | Bloody sputum; Change in caught or a new cough; Chronic, unexplained cough > 2 weeks; Lower respiratory illness (i.e. cough, shortness of breath, pneumonia); Shortness of breath |
| Upper gastrointestinal tract | UGT | Nausea/Vomiting; New mouth sore; Sore throat; Vomiting |
| Lower gastrointestinal tract | LGT | Diarrhea; Pain in the abdomen or rectum |
| Central nervous system | CNS | Severe headache; Stiff neck |
| Peripheral nervous system | PNS | Anosmia (lack or alteration of smell); Dysgeusia (lack or alteration of taste); Muscle pain; New onset of pain |
| Genitourinary system | GUS | Burning or pain with urination; Increased urination; Unusual genitourinary discharge or irritation; Vaginal pus; Delay in uterus reduction |

### Data analysis

Data set was analyzed in LibreOffice spreadsheets and Python 3. Specific statistical methods are stated with the presented results.

## Results and discussion

### General patient description

The analysis of underlying conditions, clinical outcomes, and symptoms were performed over 10,305 adult patients that were confirmed positive for COVID-19 and were hospitalized in Houston Methodist Hospital (HM) from the beginning of the pandemics till the April of 2021. Table 2 provides detailed description of patient demographics, social habits, comorbidities, and critical hospitalization events, like ICU care, ventilation, and the outcomes. 89.9% of patients survived COVID-19 infection. No racial or ethnic differences were found between the surviving and deceased cohorts. All hospitalized patients were obese and elderly on average. The group of deceased patients is dominated by older male patients who have slightly lower BMI (30.4 vs 31.7 in the alive group). Drug and tobacco use were no different among both groups of patients, however alcohol use was slightly lower in the group of deceased patients. Comorbidities, provided by Charlson Comorbidity Index (CCI), were statistically significantly more pronounced in the deceased patients across the most of comorbidities, except AIDS-HIV. While most of all hospitalized patients needed oxygen, the deceased patients had substantially higher need for ventilation (x10 times) and ICU care (x6 times). The hospitalization and ICU stay were also substantially longer among the deceased patients. The deceased patients also had lower vaccination rate compared to the hospitalized patients surviving the infection.

**Table 2.** Hospitalized COVID-19 patient information. Significance calculated by t-test and proportion tests between alive and deceased patient cohorts: ^*^ - p < 0.05, ^**^ - p < 0.01, ^***^ - p < 0.001.

| Characteristics | All<br>(n=10305) | Alive<br>(n=9263) | Deceased<br>(n=1042) | Significance |
| --- | --- | --- | --- | --- |
| **<u>Demographics</u>** |  |  |  |  |
| Gender (Male) | 53.3% | 52.4% | 61.4% | *** |
| Age (years) | 59.7 ± 16.2 | 58.5 ± 16.1 | 69.7 ± 12.8 | *** |
| BMI | 31.6 ± 8.0 | 31.7 ± 8.1 | 30.4 ± 7.9 | *** |
| Ethnicity (Hispanic) | 36.6% | 36.7% | 35.4% |  |
| Race (Caucasian) | 67.9% | 67.8% | 69.2% |  |
| Race (Afro-American) | 19.5% | 19.7% | 17.8% |  |
| Race (Native American) | 1.1% | 1.1% | 1.2% |  |
| Race (Asian) | 5.9% | 5.9% | 6.0% |  |
| **<u>Social habits</u>** |  |  |  |  |
| Alcohol | 25.8% | 26.4% | 20.3% | *** |
| Drugs | 1.4% | 1.4% | 1.1% |  |
| Tobacco | 4.3% | 4.3% | 4.0% |  |
| **<u>Charlson Comorbidity Index</u>** |  |  |  |  |
| Myocardial Infarction | 16.9% | 14.2% | 41.6% | *** |
| Congestive Heart Failure | 19.2% | 16.7% | 40.8% | *** |
| Peripheral Vascular Disease | 14.2% | 13.1% | 24.4% | *** |
| Cerebrovascular Disease | 14.4% | 13.0% | 26.9% | *** |
| Dementia | 7.4% | 6.7% | 13.9% | *** |
| Chronic Pulmonary Disease | 24.2% | 23.5% | 30.7% | *** |
| Connective Tissue Disease-Rheumatic Disease | 3.8% | 3.6% | 5.3% | ** |
| Peptic Ulcer Disease | 3.0% | 2.8% | 4.9% | *** |
| Mild Liver Disease | 10.3% | 10.0% | 12.4% | ** |
| Diabetes without complications | 44.7% | 43.1% | 59.1% | *** |
| Diabetes with complications | 18.4% | 16.8% | 32.7% | *** |
| Paraplegia and Hemiplegia | 2.4% | 2.1% | 5.0% | *** |
| Renal Disease | 24.7% | 22.4% | 44.9% | *** |
| Cancer | 8.7% | 8.3% | 12.7% | *** |
| Moderate or Severe Liver Disease | 1.7% | 1.4% | 3.8% | *** |
| Metastatic Carcinoma | 6.6% | 6.3% | 9.6% | *** |
| AIDS-HIV | 0.6% | 0.6% | 0.4% |  |
| **<u>Hospitalization events</u>** |  |  |  |  |
| O<sub>2</sub> | 84.7% | 83.1% | 98.7% | *** |
| Ventilation | 15.0% | 7.9% | 77.8% | *** |
| ICU | 78.8% | 14.4% | 81.7% | *** |
| ICU stay (days) | 2.7 ± 8.3 | 1.4 ± 5.6 | 13.9 ± 16.0 | *** |
| Hospitalization stay (days) | 7.5 ± 8.6 | 6.5 ± 6.7 | 17.0 ± 15.3 | *** |
| COVID19 vaccinated | 18.0% | 19.8% | 1.9% | *** |
| Outcome (alive) | 89.9% | 100.0% | 0.0% | *** |

The length of hospitalization was very different among surviving and deceased patients. The patients surviving the infection were hospitalized on average up to 7 days, while the stay of deceased patients exceeded two weeks (Figure 1A). Age distributions among both patient groups revealed deceased patient are above 50 years old; this group had no patients younger than 40 years (Figure 1B). The age of patients above 90 was capped at 90 for deidentification purposes forming the corresponding peak in the age distribution plots. Despite differences in hospitalization stay and age, both patient groups show similar BMI distributions (Figure 1C). BMI indicated obesity spread across all patient groups exacerbated by cardiovascular and diabetic comorbidities (Table 1), which is specifically strongly expressed among the deceased patients.

**Figure 1.**
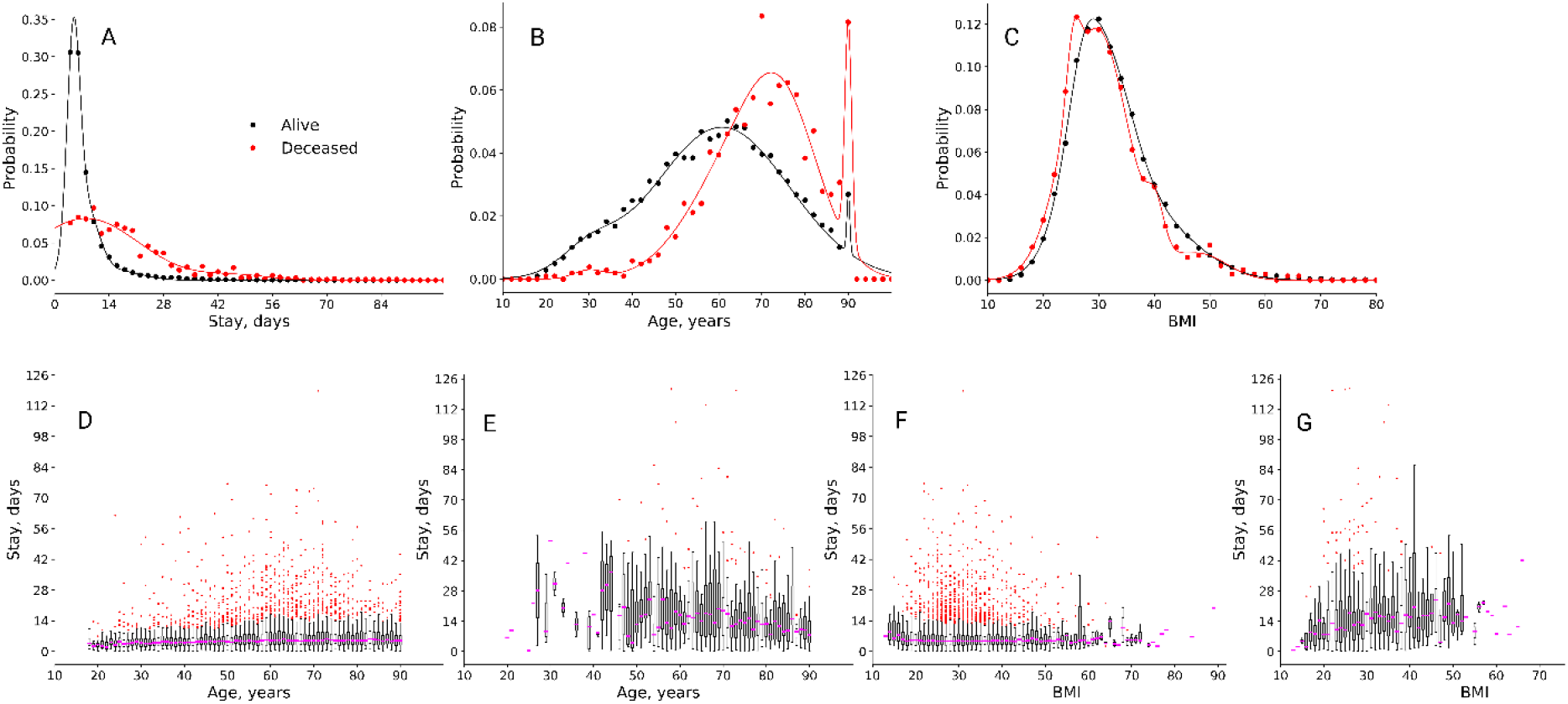
Hospitalization length of COVID-19 patients and its relation to patient age and BMI. A – Hospitalization distribution among alive and deceased patients. B – Age distribution among alive and deceased patients. C – BMI distribution among alive and deceased patients. D and E – Length of hospitalization based on age of alive and deceased patients, respectively. F and G - Length of hospitalization based on BMI of alive and deceased patients, respectively.

The dependence of hospitalization stay on age was analyzed by boxplots (Figure 1D - alive, E - deceased). Patient groups are dominated by 7 and 14 day stay for alive and deceased patients, respectively. Both groups show many outliers, especially in the age groups of 50 to 70 years. Hospitalization stay dependence on BMI was similar to age (Figure 1F - alive, G - deceased), although the trend of stay increased for BMI > 20 in the group of deceased patients could be noticed.

### Symptomatic burden

Symptoms and their corresponding recording time were extracted from patient flow sheets. The hospital admission time served as a reference and is set to zero. 40 unique symptoms were grouped based on their anatomic location to simplify our analyses and improve statistics as it is described in Methods. Table 3 tabulates symptoms recorded in all, alive, and deceased patients. An absolute majority of patients had experienced some sort of symptoms during their hospitalization. The most pronounced symptoms were associated with Lower Respiratory System (LRS) and Systemic (SYS) based on the symptom abundance among patients and the number of recorded symptoms per patient. Deceased patients had larger symptomatic burden, especially in SYS, Upper Respiratory System (URS), LRS, Lower Gastrointestinal Tract (LGT), and Genitourinary System (GUS), as well as overall symptomatic burden. However, patient surviving COVID-19 infection experienced more symptoms associated with Central Nervous System (CNS) and peripheral Nervous System (PNC).

**Table 3.** Symptomatic burden in patients. Results are presented as a fraction of patients experiencing symptoms (%) and average number of records of specific symptoms per patient (± standard deviation). Abbreviations: SYS – Systemic, URS – Upper Respiratory System, LRS – Lower Respiratory System, UGT – Upper Gastrointestinal System, LGT – Lower Gastrointestinal System, CNS – Central Nervous System, PNS – Peripheral Nervous System, GUS - Genitourinary System. Significance: ^*^ - p < 0.05, ^**^ - p < 0.01, ^***^ - p < 0.001.

| Symptoms | All (n=10305) |  | Alive (n=9263) |  | Deceased (n=1042) |  | Significance |
| --- | --- | --- | --- | --- | --- | --- | --- |
|  | % | n | % | n | % | n |  |
| All | 94.1% | 12.3 ± 13.4 | 93.6% | 10.7 ± 11.2 | 97.1% | 26.3 ± 21.3 | *** |
| SYS | 47.2% | 1.3 ± 2.6 | 46.2% | 1.2 ± 2.3 | 56.4% | 2.5 ± 4.1 | *** |
| URS | 3.6% | 0.1 ± 0.3 | 3.3% | 0.0 ± 0.3 | 5.9% | 0.1 ± 0.3 | ** |
| LRS | 86.9% | 8.8 ± 9.9 | 85.9% | 7.7 ± 8.4 | 94.9% | 18.8 ± 14.9 | *** |
| UGT | 17.5% | 0.3 ± 0.9 | 17.8% | 0.3 ± 0.9 | 14.7% | 0.3 ± 0.8 | |
| LGT | 12.9% | 0.4 ± 1.4 | 12.4% | 0.3 ± 1.4 | 16.8% | 0.4 ± 1.4 | * |
| CNS | 5.8% | 0.1 ± 0.3 | 6.1% | 0.1 ± 0.3 | 3.6% | 0.0 ± 0.2 | ** |
| PNS | 14.2% | 0.2 ± 0.5 | 14.9% | 0.2 ± 0.5 | 8.2% | 0.1 ± 0.4 | *** |
| GUS | 1.9% | 0.0 ± 0.3 | 1.7% | 0.0 ± 0.3 | 4.0% | 0.1 ± 0.4 | *** |

Patient symptoms were analyzed as a function of time. Symptom burden peaked within the first 12 hours after hospitalization and switched to monotonic symptom remission after 12 hours. We assumed the high rate of symptoms is observed before medication administration and before the projected time to observe any treatment effects. As it is illustrated by the evolution of all symptoms in Figure 2A, symptoms burden decreases much faster for the patients that survive the infection compared to deceased patients; similar trends were observed for SYS and LRS (Figure 2B and C). The symptom burden was mostly dominated by LRS, in all COVID-19 patients. Because symptomatic burden decreases slower than in the patients surviving COVID-19, the ratio of symptom remission rates could be perceived as a surrogate risk marker for hospitalized patients. The ratio of symptom remission rates is plotted in Figure 2D showing that morbidity risk increases exponentially with time. The critical time of 7 days is observed for most of the symptoms at which the cross-over of symptom burden kinetics is found. The morbidity risk is 3 and 8 times higher after 2 and 3 weeks, respectively.

**Figure 2.**
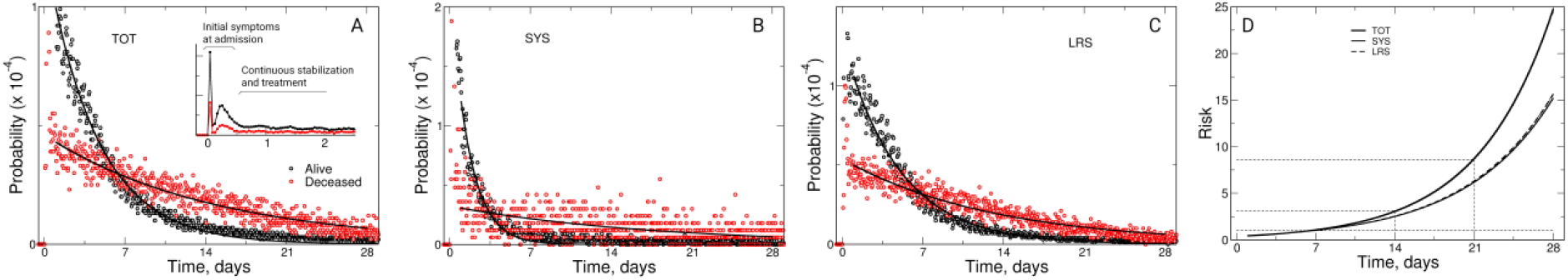
Longitudinal symptom analysis of hospitalized COVID-19 patients. A – Total symptomatic burden for alive and deceased patients. The inset shows the full probability scale and characteristic temporal segments of symptoms. All and other plots focus on symptom data 12 hours after admission. Lines represent best exponential fits. B – Temporal evolution of systemic symptoms (e.g. chills, fever). C - Temporal evolution of LRS (e.g. shortness of breath pneumonia, etc.). D – Morbidity risk as the ratio between symptom burden from alive and deceased patients from the corresponding fits in A, B, and C.

To better understand survival in relation to the symptom burden, the time of the latest recorded symptom for a specific symptom groups was analyzed as a surrogate marker for symptom endurance (Figure 3). The analysis suggests that some symptoms, such as UGT or SYS, vanished relatively fast within a couple of days on average. However, LRS symptoms lingered the longest with a persistence for up to few weeks.

**Figure 3.**
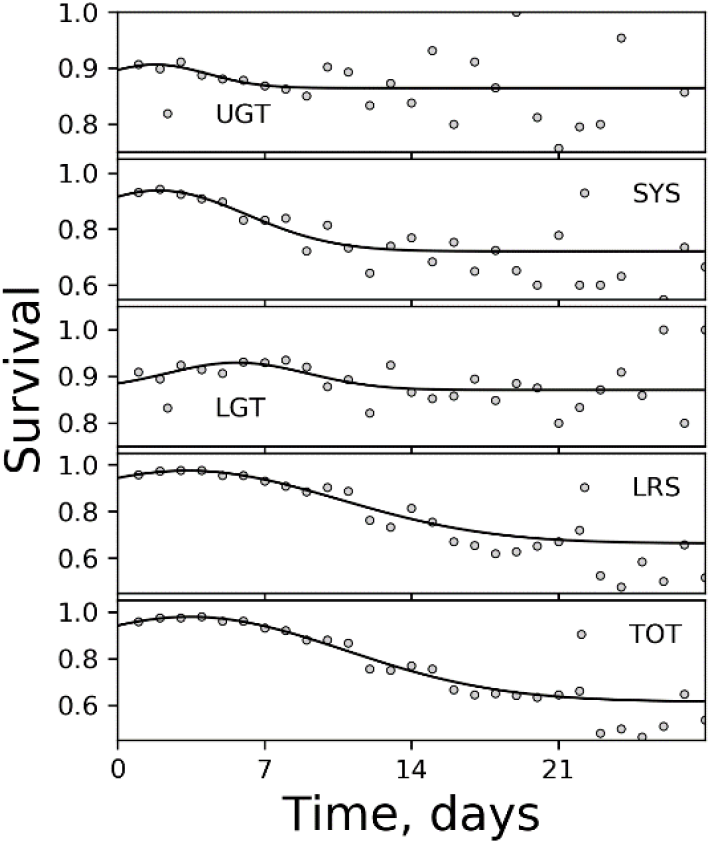
Survival of hospitalized patients for different symptom groups based on the time of the last recorded symptom for a patient. The plots are presented from fastest to slowest symptom kinetics with the normal fit approximating the kinetics of symptoms.

### Symptom remission rate

Changes in symptom burden take place over weeks taking the shape of exponential decay. We have analyzed characteristic times of symptom remission by fitting their distributions with the exponential decay function to quantify how fast symptom changes take place among alive and deceased patients (see *Supplemental Material*). We have found that the first three days can be a good approximation of the whole symptom remission dynamics over many weeks by finding positive correlation between characteristic times (from exponential fitting) and linear rates during the first 3 days with R^2^ = 0.79 and 0.88 for alive and deceased patients, respectively.

Because the symptom phase over the first few days correlates directly with the symptom kinetics over the whole duration of hospitalization, we calculated linear symptom remission rate for individual patients with 5 and more symptoms recorded within the 1^st^ weeks of hospitalization. This approach tests if information obtained on the level of whole cohort is translatable to an individual patient. Furthermore, an individual rate of symptom remission can be potentially used as a prognostic tool to help forecast patient hospitalization and clinical outcome outlooks. The distribution of the symptom remission rate was gaussian-like and centered around zero for deceased patients. Figure 4A shows the distribution of symptom remission rate based on all recorded symptoms. The patients surviving the infection had bi-modal gaussian distribution with a second gaussian peak positioned at -0.5 symptom/day. The bi-modal distribution was separated by the minimum at -0.25 symptom/day. Thus, we have designated two groups: first, the patients with fast symptom remission with < -0.25 symptom/day and second, the patients with slow symptom remission with > 0.25 symptom/day. The hospitalization of patients was the longest for the patients characterized with lack of dynamics in symptoms during the 1^st^ week of their hospitalization (Figure 4 B,C). The hospitalization length was shorter for the patients with fast symptom remission and the average hospitalization stay increased for surviving patients with slower symptom remission.

**Figure 4.**
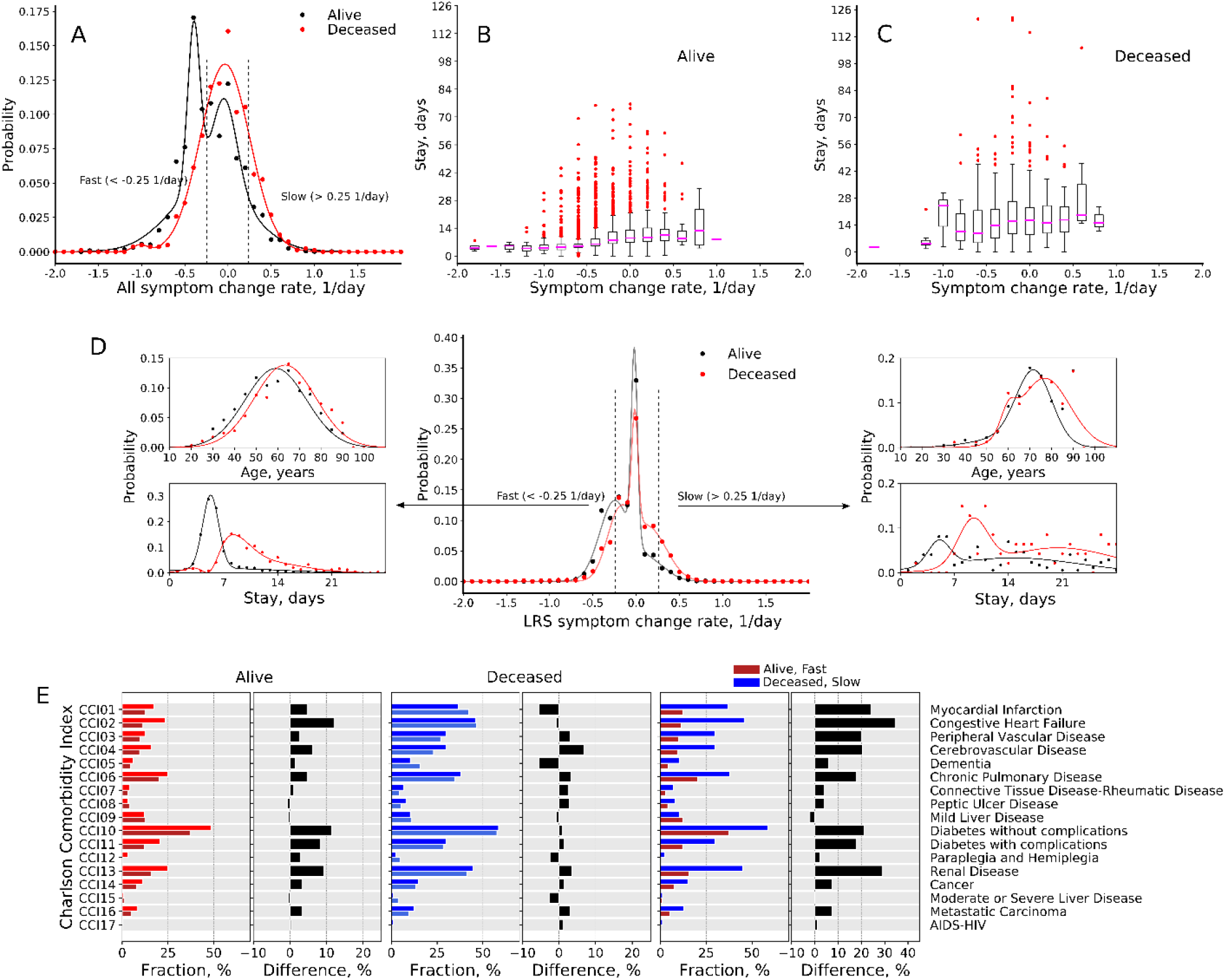
Symptom remission rate of the COVID-19 patients during the first week of hospitalization. **A** – The rate distribution for alive and deceased patients. **B** and **C** – Hospital stay based on symptom remission rate for alive and deceased patients, respectively. **D** – Comparison of comorbidities among alive and deceased patients with slow and fast remission rates.

We analyzed symptom remission rates for LRS symptoms as they are the most prevalent and frequent symptoms among all hospitalized COVID-19 patients (Figure 4D). The symptom remission rates of alive and deceased patients have a peak at zero with side-shoulders expressed differently in alive and deceased patients. Patients with fast (< -0.25 symptom/day) and slow (> 0.25 symptom/day) remission rates had similar BMI. However, differences were noted in their age and hospitalization stay based on remission rates. Patient age with fast symptom remission were 60 and 65 years for survived and deceased patients, respectively, and those patients were younger than the patients with slow symptom change rate, 70 and 75 years, respectively. Hospitalization stay was also longer for the patients with slow remission rates, as well as deceased patients in comparison to the surviving patients.

Comorbidity comparison of patients with LRS < -0.25 and > 0.25 symptom/day is presented in Figure 4D. All hospitalized patient had a significant number of comorbidities. However, the deceased patients had twice more comorbidities on average than patients surviving COVID-19. Among alive patients the fast symptom remission rates were related to less comorbidities. Comorbidities related to various cardiovascular diseases, diabetes, and renal diseases were associates with slower symptom remission in alive patients. Deceased patients show almost twice more comorbidities present and differences between patient with slow and fast remission were smaller. The largest difference was noticed comparing alive patients with fast symptom remission against deceased patients with slow symptom remission. Again, the major differences were associated with cardiovascular diseases, diabetes, and renal diseases.

## Discussion

The results of our analysis indicated that hospitalized patients are dominated by symptoms in LRS (e.g. shortness of breath, pneumonia) and SYS (e.g. fever) groups. Deceased patients experienced more symptoms than the patients who survived the infections. The larger recorded number of symptoms may be also related to longer hospitalization times among deceased patients. The overall symptom dynamics was found to be similar to other studies [11, 12]: symptom burden peaks at the beginning and decreases over time. Symptoms from LRS were the most abundant among all hospitalized patients providing a good view into symptom dynamics. The symptom remission rates were faster among surviving patients within the 1^st^ week of their hospitalization compared to deceased patients. The symptom remission rates equalized at the day 7 for both groups. Similar profiles were noticed the SYS symptoms and all aggregated symptoms. Based on the symptom remission ratio between deceased and alive patients, the morbidity risk increases 3 and 8 times after 1 and 3 weeks, respectively.

The analysis of symptom remission rates within individual patients have revealed the presence of patient sub-populations with fast and slow remission rates. The analysis was done only for patients that had 5 or more symptom records within the 1^st^ week of their hospitalization withing a specific group of symptoms. The chosen time period of one week is related to the observation that remission rates intersect at approximately 7 days in hospitalization. Thus, the ability to understand the symptom patterns within early hospitalization stage, e.g. within 48 or 72 hours, may be helpful to provide meaningful prognostic information about the hospitalization outlooks for an individual patient. The patients with fast symptom remission have shown shorter hospitalization, which is also related to increased chances of survival. Deceased patients and patients with longer hospitalization had more comorbidities, especially cardio-vascular diseases, diabetes, and renal diseases. Cardiovascular and diabetic comorbidities were also found associated with higher risks of death in another study [16]. Although, all hospitalized patients had significant comorbidity burden despite their clinical outcomes. Patient BMI comparison suggested that all hospitalized patients had various degrees of obesity and BMI was not correlating to clinical outcomes. Obesity was already noticed to correlate with higher prevalence of viral infections [17, 18]. Patient age was found to be an important feature where older age was correlating to longer hospitalization and higher mortality. Among deceased patients, there were almost no patients of age 40 and younger.

An additional view into survival was provided by analyzing symptoms based on their last recorded time. Specific symptom groups have shown symptom ending at different times: symptoms ended in patients in increasing order: UGT<SYS<LGT<LRS<TOT (Figure 3). The analyzed fraction of survived patient as a function of hospitalization time has relatively narrow window for UGT or LGT symptoms, which was ∼10% and comparable to the overall patient survival. However, SYS, LRS, and TOT symptoms showed 30-40% dynamic range, which makes them useful for prognostic purposes.

One of the strengths of our study is that we have simplified symptom analysis by grouping them based on anatomical location associated with symptoms. The basis for this is that once the infection spreads through organs, e.g. lungs or intestines, there is no single unique symptom but rather a set of them. For example, diarrhea and abdominal pain may both be related to lower gastrointestinal tract. This approach also reduces the number of variables, improves the statistics, and provides a clearer picture of infection dynamics. But it necessary to acknowledge several critical aspects of symptom analysis. One of them is that symptom collection may be inconsistently elicited form patients by clinicians and patients may be monitored differently because of clinical needs. Furthermore, the sensitivity and specificity of symptomatic analysis may be not be adequate at the current stage because the data collection inconsistencies mentioned above.

Longitudinal symptoms dynamics holds a lot of analytical depth, because the temporal aspect of how symptom burden changes reflects the patient health. The number of longitudinal studies on COVID-19 symptoms is very limited to date. During peaks of COVID-19 pandemics hospitals face challenges from operational and care perspectives, where planning of resources and personnel become critical. Being able to predict or forecast even basic events such as hospitalization length or predict outcome events based on early symptoms may be useful in developing pre-planning procedures to make sure that patients will receive the best suitable care and have the best outcomes. Furthermore, symptom recording, and analysis does not require sophisticated equipment and may be applied in various hospitalization environments.

Our findings indicate that symptom analysis can hold prognostic power. While the overall finding about symptoms may not be new, such an analysis and results have not been reported before. The symptom collection does not rely on sophisticated equipment and does not requires complex data collection. It is worth mentioning that symptom collection depends on nurses and workload, which can be intense during pandemics like COVID-19. Data sparsity should be expected in these situations. Therefore, improving symptom data collection may bring more economic gratitude to the healthcare industry by enabling to increase healthcare personnel efficiency and optimize patient care at the organizational level. This study relied on retrospective symptom data collected through normal hospital operation and data collection was not optimized for any specific purpose. Despite that, our results suggest that having a dedicated symptom collection and analysis may lead to even better prognostic information which could be individually tuned for each patient. The dynamics of symptoms within the first days of their hospitalizations should be the most useful period for data collection by approaching the recording on more consistent bases or its higher frequency. While this study analyzed the symptoms and their wear-off rates, and their relations to various patient features, the application of Deep Learning methods will be the next stage for symptom analysis to create versatile prognostic models.

## Conclusion

The possible relations of symptoms and hospitalization are known in general, but were not analyzed in the hospitalized COVID-19 patients to date. The analysis of hospitalized COVID-19 patients in the Houston Methodist Hospital at the Texas Medical Center have revealed that longitudinal symptom analysis may possess prognostic potential regarding patient hospitalization stay and clinical outcomes. LRS symptoms were the most abundant among patients and could be used as the main data source, but the SYS and TOT symptoms can be used as well. The hospitalized patients were obese on average and possessed significant number of comorbidities. A lower chance of patient survival and longer hospitalization was associated with older patients, slow symptom wear-off rate, and comorbidities like cardio-vascular diseases, diabetes, and renal diseases. Improved and consistent system reporting should improve the prognostic capabilities of longitudinal symptom analysis. This technique could be applied with little investments and high return to healthcare operations in various healthcare setting agnostic to technological excellence. While symptom analysis reliability can be clouded by lack of sensitivity, confounding factors, and inconsistent symptom reporting, Machine Learning and Artificial Intelligence tools may help to mitigate those shortcomings.

## Data Availability

All data produced in the present study are available upon reasonable request to the authors. The original data can be requested only from Houston Methodist Hospital.

## Acknowledgments

We thank Stephen L. Jones, MD MSHI (HMH) for valuable discussion contributing to the success of this study.

## Data availability

The raw data sets can be requested by contacting HMH. The data sets generated in this study for the purpose of tables and figures can be requested directly form the authors.

